# USES OF THE SKEW-LOGISTIC FUNCTION FOR MULTI-WAVE FUNCTIONS

**DOI:** 10.1101/2022.12.19.22283694

**Authors:** Russell Cheng, Brian Williams

**Affiliations:** University of Southampton, Highfield, SO17 5BJ, United Kingdom; SACEMA, Stellenbosch University, South Africa

**Keywords:** Covid-19 waves, climate change, carbon negative

## Abstract

The Skew-Logistic (SL) function has been proposed to model a real-life dynamic process which rises monotonically to a peak followed by a monotonic decline. It was introduced to model the first stage of the Covid-19 pandemic to forecast the behaviour of Covid. Then, with different controls and variants, Covid levels rose and fell in what might be called a Multi-Wave (MW) behaviour; with the waves not necessarily the same size. This paper shows how using the SL function for one wave can be modified to model the MW situation. We apply it to two examples. One is to Covid -19, to examine its most recent behaviour. The other is to climate change, the most serious issue of our time. Ensuring that the world simply achieves carbon-equality is not enough. We have to rapidly achieve carbon-negativity to prevent bringing an end to the world as we know it.

## 1 INTRODUCTION

The Skew-Logistic (SL) function was introduced in Dye *et al*. (2020) to compare the first wave of the Covid-19 epidemics in different European countries. The mathematical form is described in the Supplemental Materials of the paper, which also describes how the model can be fitted to real data using standard statistical techniques. This paper shows how the method can be used to firstly fit to the individual waves of a sample of Multi-Wave (MW) data and how all these individual waves can be then combined to create an overall simultaneous fit the entire data sample. It will be convenient to summarise the single wave fitting first, which is done in Section 2. Then, in Section 3, we discuss the MW fitting method.

Our first application is described in Section 4 where we fit the Covid-19 Active Cases that occurred in the United Kingdom from March 2022 till November 2022 covering the three most recent waves.

In our second application we fit the SL-MW model to the annual atmospheric carbon dioxide level, in parts per million (CO_2_ ppm) from 1800 to 2022. We also fit the model to the world average Temperature (^0^C). We then use the fits to predict the CO_2_ and Temperature levels until 2100.

## 2 SINGLE WAVES

Fitting the SL function to the observation of one wave of the process of interest forms the basis for fitting several SL functions to MW data. We therefore start by summarising the description of the SL function given in Dye et al. (2020). The SL function takes the form:

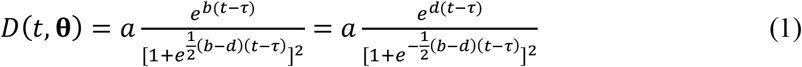

where *D*(*t*,**θ**) is the prevalence or incidence of the quantity of interest, this being the number of active Covid-19 cases in Example 1 and CO_2_ in Example 2. All four parameters are readily interpretable. The parameter *a*, is close to the maximum value of *D*(*t*), whilst *τ* indicates the location of the maximum. The parameters *b* and *d*, respectively indicate the rates of rise and decline of *D*(*t*). As shown in Equation 1, *b* and *d* are mathematically identical. We write **θ** = (*σ, a, b, d, τ*) where *σ* is the standard deviation of an individual observation. As dependence of *D*(*t*) on the parameters is obvious, **θ** is usually omitted.

We assume that *b* > 0 and *d* < 0 so that the term (*b* − *d*)/2 in the denominator is always positive. *D*(*t*) ≅ *e*^*b*(*t*∼*τ*)^ as *t* → −∞, so that *b* gives the exponential rate of increase of *D*(*t*) as *t* increases from −∞ and *D*(*t*) ≅ *e*^*d*(*t*∼*τ*)^ as *t* → ∞, so that *d* < 0 gives the exponential rate of decrease. Either form of *D*(*t*) in Equation 1 can thus be used when fitting to data, with the signs of *b* and *d* showing which role they have.

The explicit maximum value of *D*(*t*) is

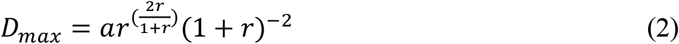

where *r* = −*b*/*d*. The maximum is at

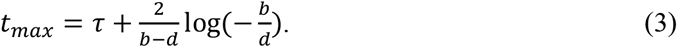

Equations (2) and (3) show that, when *b* and – *d* are close in value, *a* will be close to the maximum of *D*, and *τ* wil be close to the maximum point.

As described in Dye *et al*. (2020), the SL function can be fitted to data using the method of maximum likelihood (ML) using Nelder-Mead search. This latter needs initial parameter values to be provided. This is not always straightforward, especially with multimodal functions, particularly when the number of parameters is large. A very attractive feature with the SL function is that, its readily interpretable parameters enables the user to intervene and interactively select appropriate initial parameters are close the best. This enables Nelder-Mead to reliably obtain an accurate optimum. We can simply visually examine the data to obtain an initial estimate of its position and size. The rates of increase and decrease can then be roughly estimated to give starting values for *b* and *d*. An initial value for *a* be obtained using Equation (2), whilst the observed maximum and Equation (3) can be used to give the initial value of *τ*. In applying ML optimization, we include an extra parameter *σ*, the standard deviation of the observational error. Estimation of *σ* is included in the ML method, so that the quality of the fit is also assessed. We turn now to the MW situation

## 3 MULTI-WAVE

### 3.1 Function Formula and Data Examples

We assume a simple additive functional form when there are *N* waves, namely:

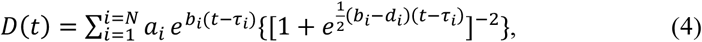

where typically, but not invariably, the *τ*_*i*_ are in increasing order *τ*_1_ < *τ*_2_ … < *τ*_N_ indicating the (approximate) location of each of the maxima of each of the waves. We give two data samples where *D*(*t*) might be fitted. The first, Example 1, comprises the observed Daily New Cases of Covid-19 from 3 March 2022 to 17 November 2022 as shown in Figure 1.

**Figure 1.**
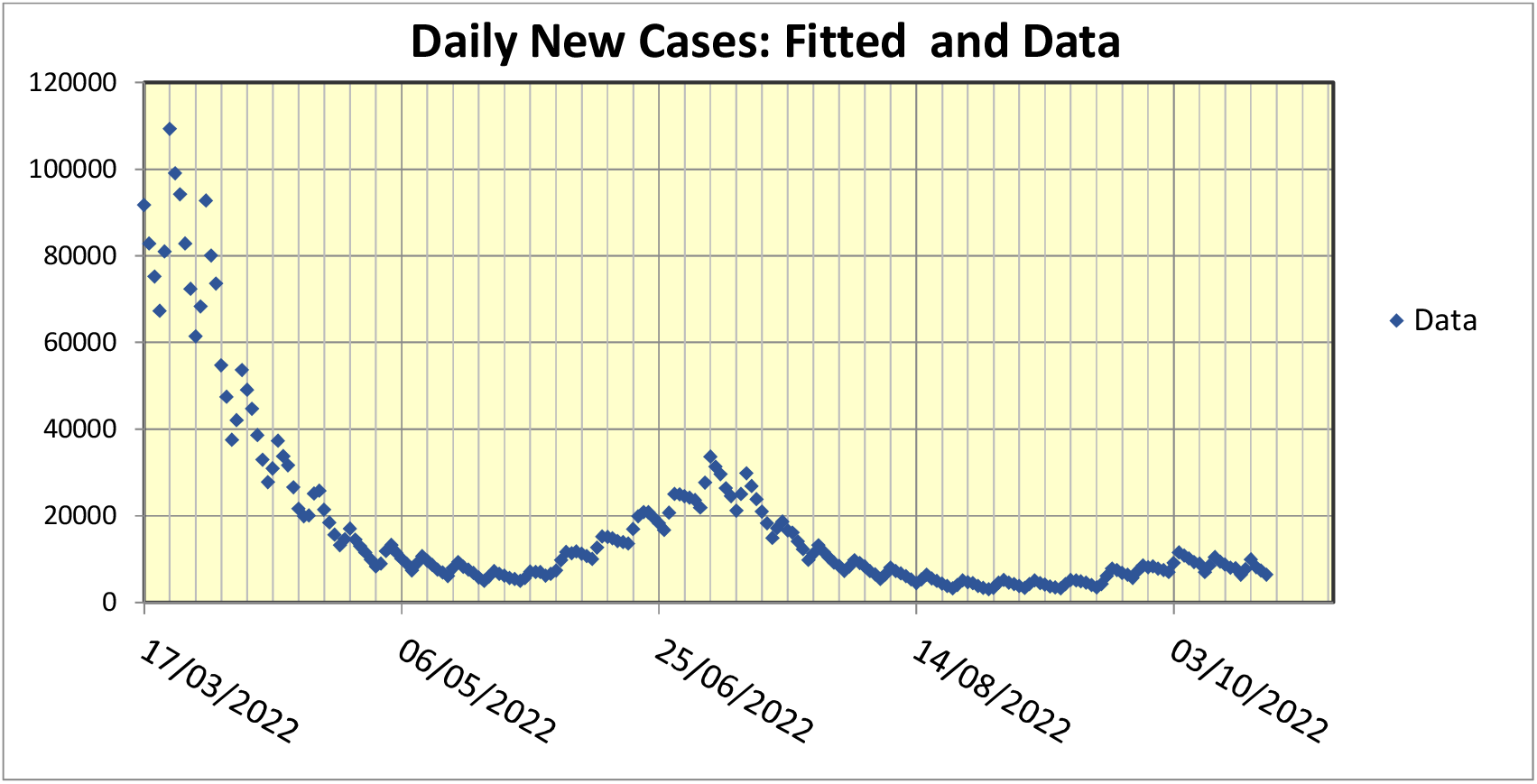
Covid-19 observations from 17 May to 16 November 2022

Example 2, comprises the observations of the global CO_2_ level from 1800 to 2022. A plot of this sample is shown in Figure 2.

**Figure 2.**
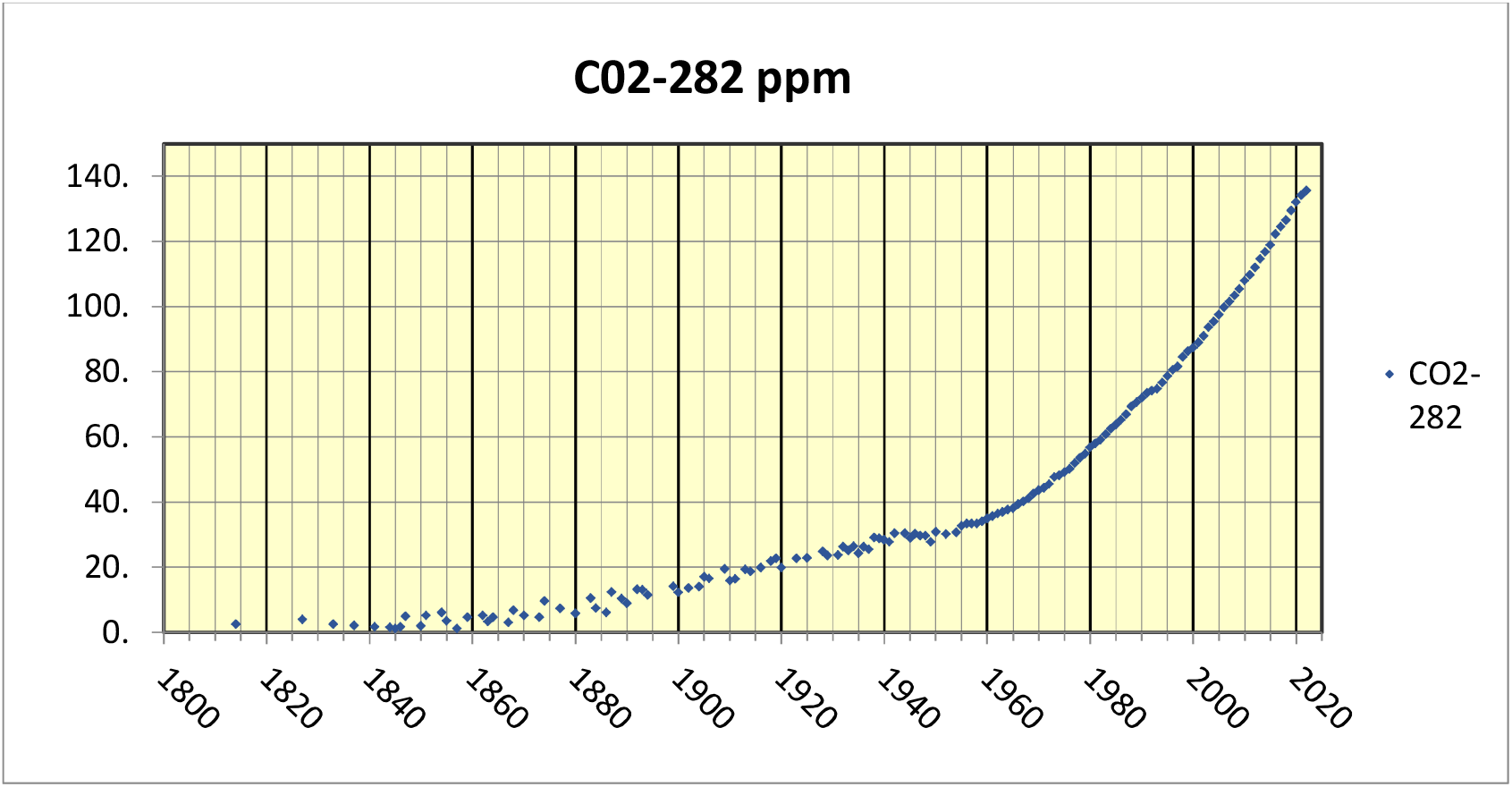
CO_2_ –282 ppm Level from 1800 to 2022

There is a noticeable peak in the sample plot where a wave is particularly prominent but this depends on the juxtaposition of the waves and where a wave peaks may not be obvious from the full sample plot. In Example 1, one might conjecture that there are three waves. In Example 2, the number of waves is not so clear and there could be one or two waves.

We consider our proposed fitting method next.

### 3.2 The Basic Method

The proposed method of fitting *D*(*t*) to data takes two stages. In the first stage the data is grouped into separate sections, with each representing the position where the data are most dependent on a particular wave. The division process could be made automatic, but if speed is not important, as is the case in our two examples, then it is simplest for the user to carry out this process by eye. As will be seen, in Example 1, the number of waves looks likely to be three. As previously intimated, the choice is less obvious in Example 2. However the initial slope variation suggests an earlier wave that becomes dominated by a second wave. Here more adjustment is needed to fix the position of the first wave and our final choice turns out to be two waves.

To find the maximum when using search methods like Nelder-Mead, a good choice of initial parameters is extremely important, particularly when the parameter count is high and the search domain is high dimensional. The problem is particularly demanding when the function is multimodal as is the case in our problem. In our case, visual evidence of a good fit of a regression line to data, together with confidence level assessment, provides reassurance that our method is satisfactory. In the next section we study the examples in more detail

## 4 FITTING METHOD

### 4.1 Covid-19 Example

The fitting problem is amenable to being tackled visually and user interaction. We indicate the general procedure which is very flexible. There are no hard and fast rules and, depending on the data, adjustments are easy to make. How a simple overall approach is to divide the fitting into two steps. We consider Example 1.

As the first step we make a visual inspection of the sample. This suggests choosing 3 waves with waves 1, 2 and 3 contributing to data points in the ranges *S*_1_ = {1,86}, *S*_2_ = {87,172}, *S*_3_ = (173,243}, respectively, where the sample size is *n =* 243. We then separately fit the single wave SL model of *D*(*t*), as given in Equation (1), to each of the data sets *S*_j_, *j =* 1, 2, 3. The fitting process is described in Cheng *et al*. (2020), Section 2 and in Dye *et al*. (2020), Supplementary Materials, and will not be repeated here. Figure 3 gives the results of this step.

**Figure 3.**
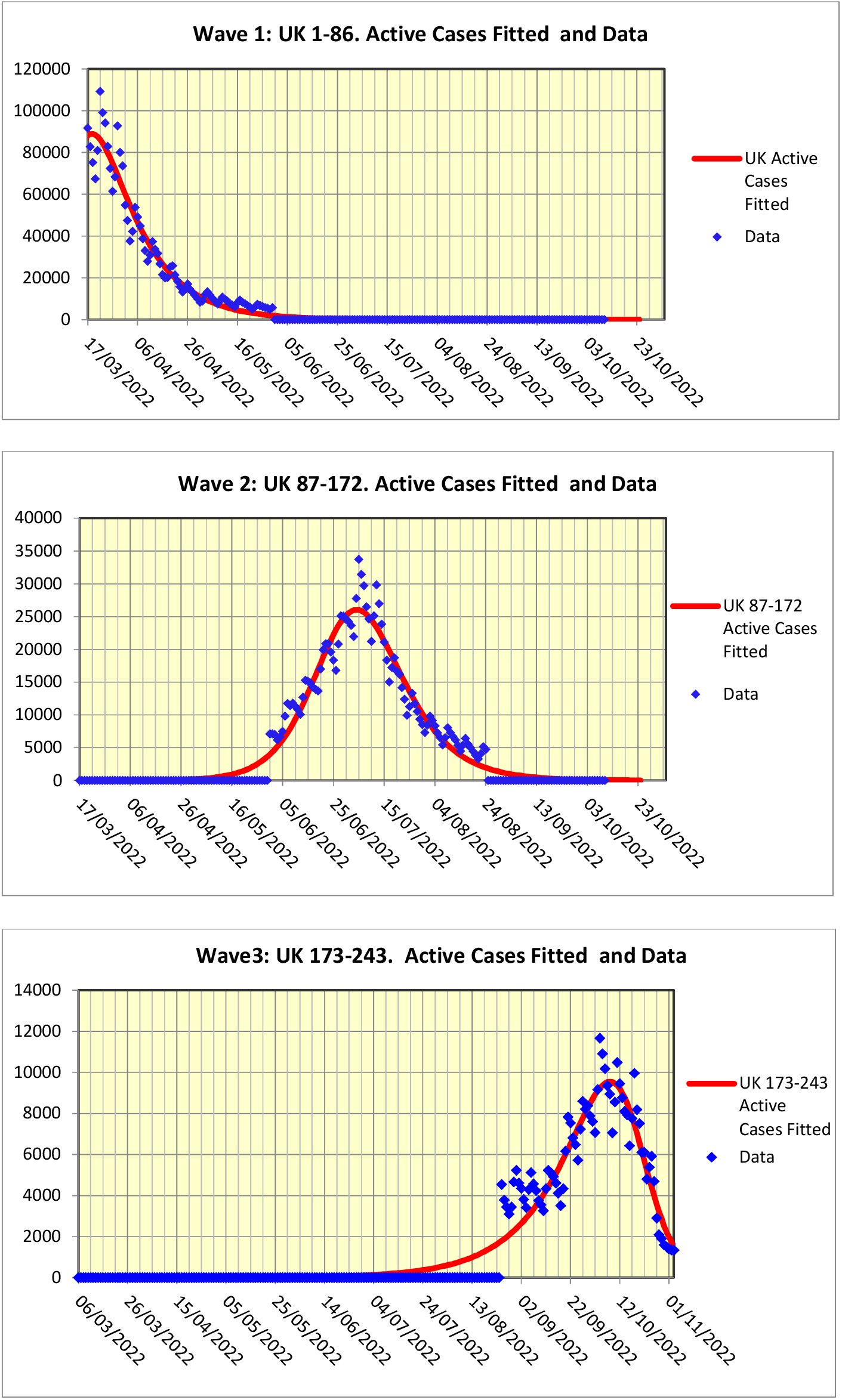
The three fitted waves for the Covid-19 sample

The fitted parameter values are given in Table 1 below.

**Table 1:** Parameter estimates obtained by dividing full sample into 3 subsamples and fitting a single wave separately to each subsample.

| Parameters | 1-86: Wave 1 | 87-172 Wave 2 | 173-243 Wave 3 |
| --- | --- | --- | --- |
| $\sigma$ | 4490 | 1630 | 748 |
| $a$ | 254000 | 101000 | 27500 |
| $b$ | 0.215 | 0.103 | 0.0492 |
| $d$ | -0.0596 | -0.0713 | -0.174 |
| $\tau$ | 44600 | 4470 | 44900 |

Once the single waves have been fitted, we proceed to the second step which is to fit the *N=*3, multiwave *D*(*t*) of Equation (4) taking θ ={*σ, a*_1_, *b*_1_, *d*_1_, *τ*_1_, *a*_2_, *b*_2_, *d*_2_, *τ*_2_, *a*_3_, *b*_3_, *d*_3_, *τ*_3_} as the vector of initial parameter values. The subscripts correspond to the number of the fitted wave. The initial multiwave *D*(*t*) obtained, using the initial parameter values of Table1, is shown (in green) in the chart of Figure 4. Note that the last segment of this curve, corresponding to wave 3, is noticeably different from the fit to wave 3 shown in the third plot of Figure 3.

**Figure 4.**
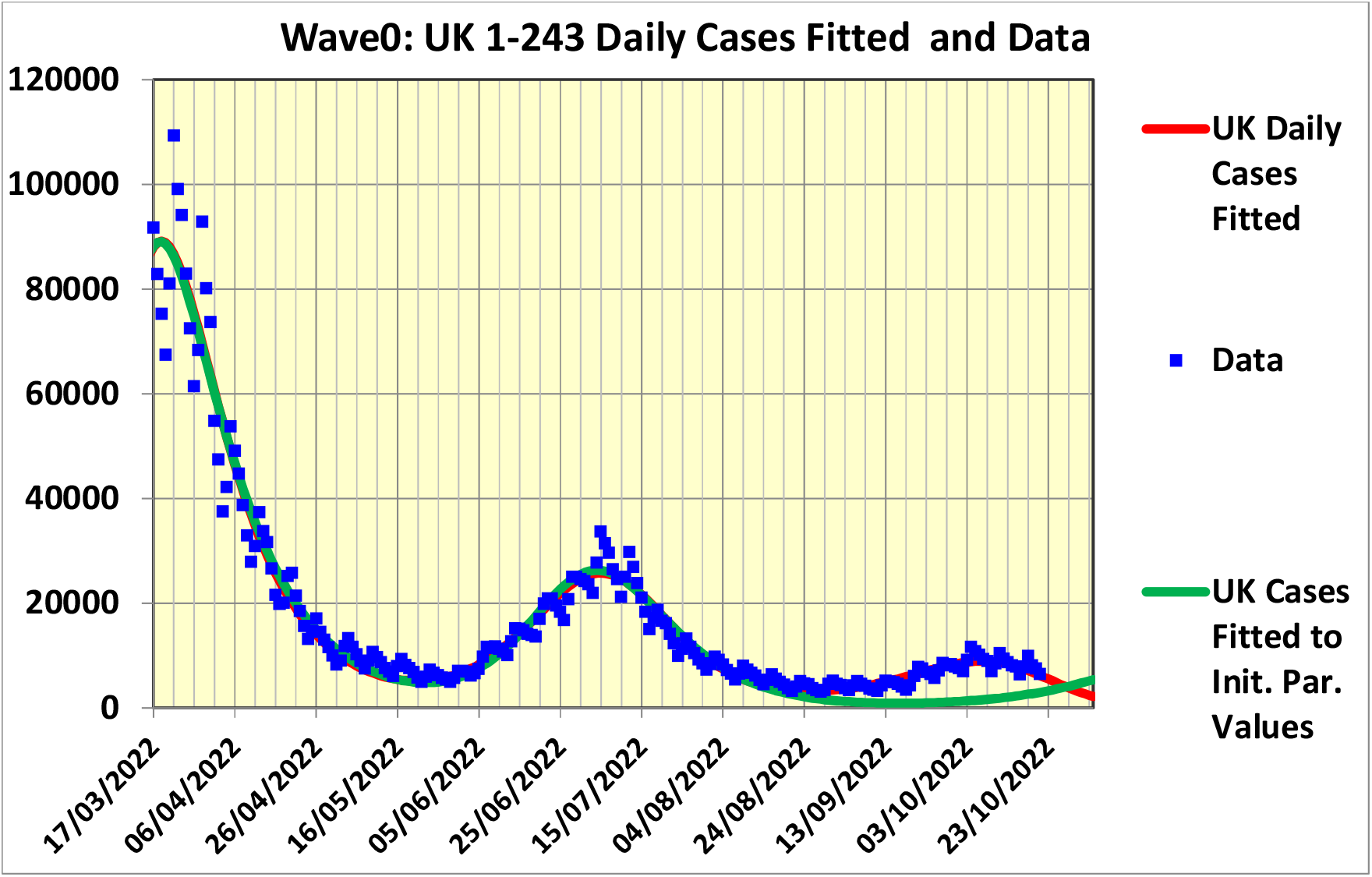
The N = 3 Multi-wave fit to the full Covid-19 sample

Figure 4 also shows the final fit (red line) obtained for the full sample, where it will be seen that the optimization greatly improves the overall fit.

The optimized parameter estimates are shown in Table 2

**Table 2:** Parameter estimates obtained by an N=3 Multi-wave to the full Covid-19 sample

| Parameters | $i = 1$ | $i = 2$ | $i = 3$ |
| --- | --- | --- | --- |
| $\sigma$ | 4610 | | |
| $a_i$ | 27000 | 102000 | 28300 |
| $b_i$ | 0.198 | 0.0817 | 0.0483 |
| $d_i$ | -0.0637 | -0.0875 | -0.135 |
| $\tau_i$ | 44600 | 4470 | 44900 |

**Table 3:** Parameter estimates obtained by dividing full GlobalCO_2_ sample into 2 subsamples and fitting a single wave separately to each subsample.

| Parameters | 1-76: Wave 1 | 77-145 Wave 2 |
| --- | --- | --- |
| $\sigma$ | 1.46 | 675.0 |
| $a$ | 111.0 | 0.0269 |
| $b$ | 0.0394 | 0.0269 |
| $d$ | -0.0196 | -0.784 |
| $\tau$ | 1939.0 | 208.0 |

### 4.2 CO_2_ Example

In this example, visual inspection of the data sample shown in Figure 2 suggests fitting two waves.

Using these single wave parameter estimates as initial values produces the *N=2* Multi-Wave fit shown in Figure 6 below.

**Figure 5.**
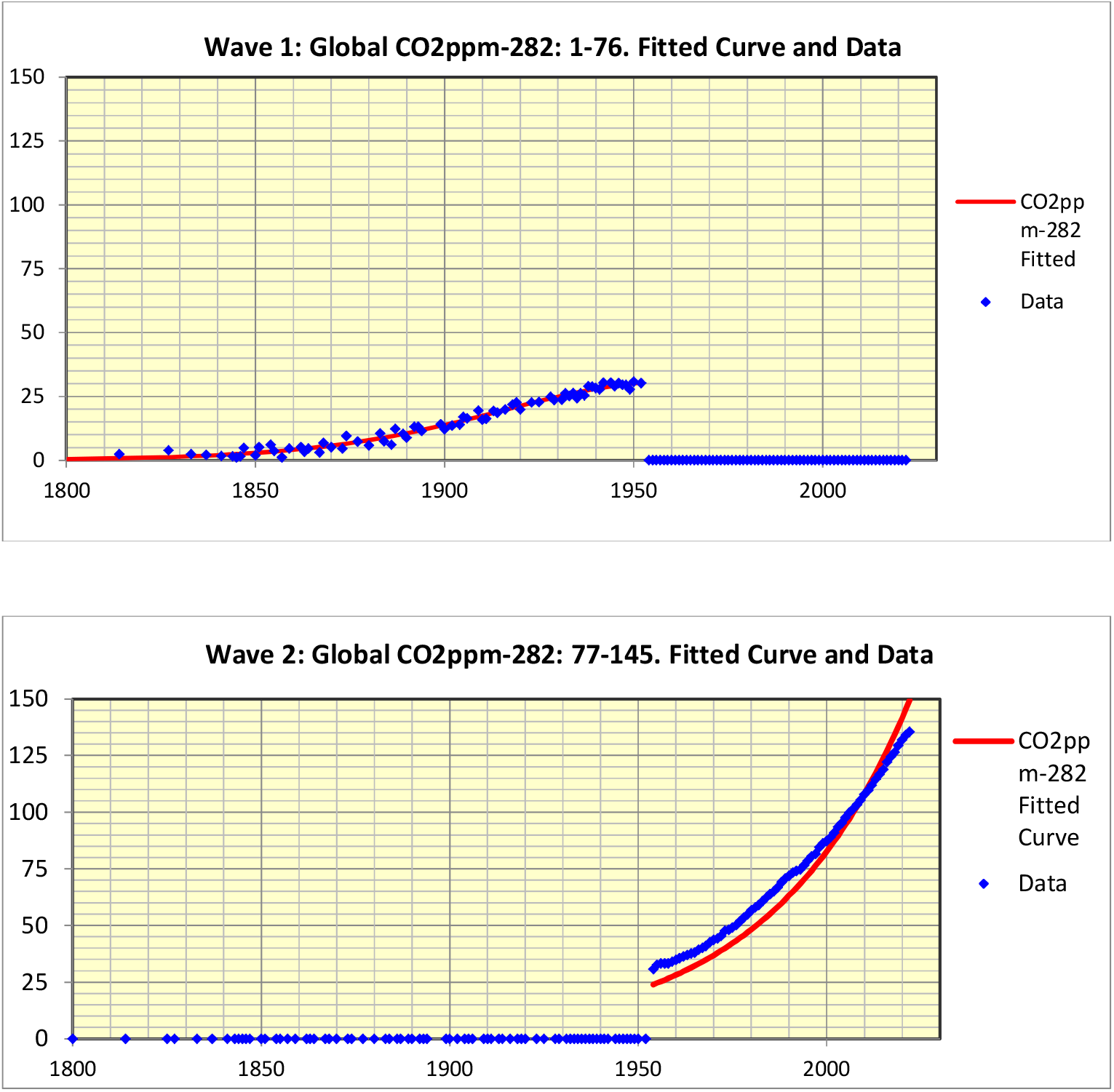
The two Single Wave fits to the Global CO_2_ ppm sample

**Figure 6(a).**
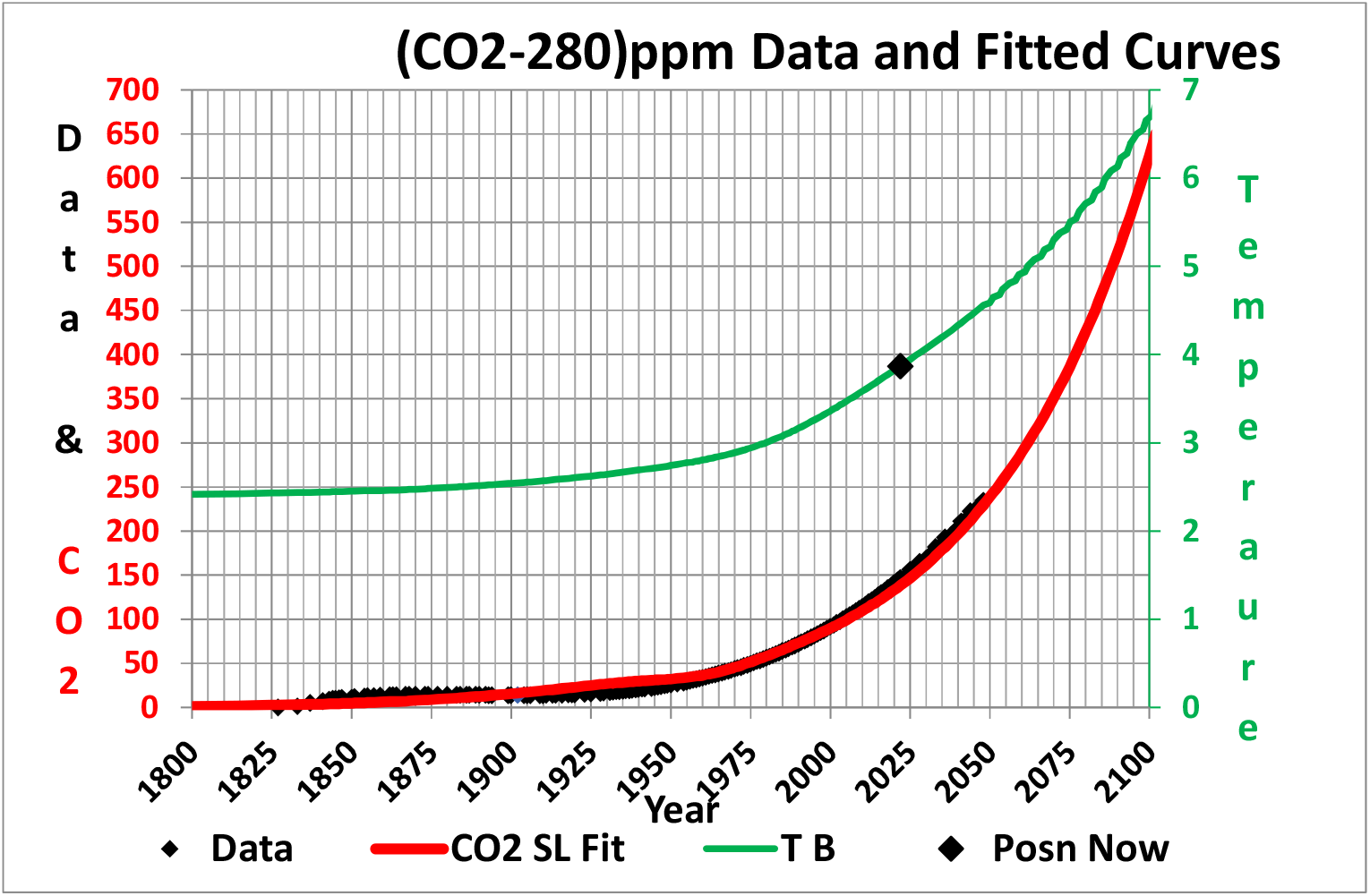
The N=2 Multi-wave fit (red, C)2 SL Fit) to the full Global CO_2_ ppm sample and the corresponding estimate of the Temperature (light green, denoted as TB) curve, calculated using a simple linear approximation with D(t) as the mantissa as described in Subsection 4.3.

### 4.3 Temperature Calculation

In this subsection we explain the calculation of the change in temperature from 1800 to 2022 that is also plotted in Figure 6. We make the simple first order linear assumption that *T*(*t*) the average global temperature in ^0^*C* is

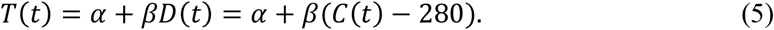

From 1800 to 2022, *C*(*t*) is global concentration level in ppm at time *t* has increased by about 48% from 280 ppm to 420 ppm, an increase of (420-280)/280 = 0.5, whilst *T*(*t*) has increased by about 1.5 degrees to 3.9 degrees. We have *C*(*t*) = *D*(*t*) + 280 so from Equation 5 we have

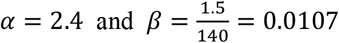

Jonas gives a formula

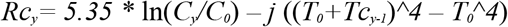

giving the between the yearly change in CO_2_ in terms of the corresponding change of year. Simply as reflection of this formula, but without using any of the considered reasoning given in (Jonas, M) we used following quartic polynomial giving the reverse calculation:

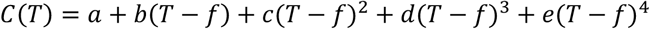

It will be seen in Figure 6(b) that the calculation gives comparable results to the other fits. The difference of the curve based on Equation (5) and the other fits arise from different base *T* values used as the initial year.

**Figure 6(b).**
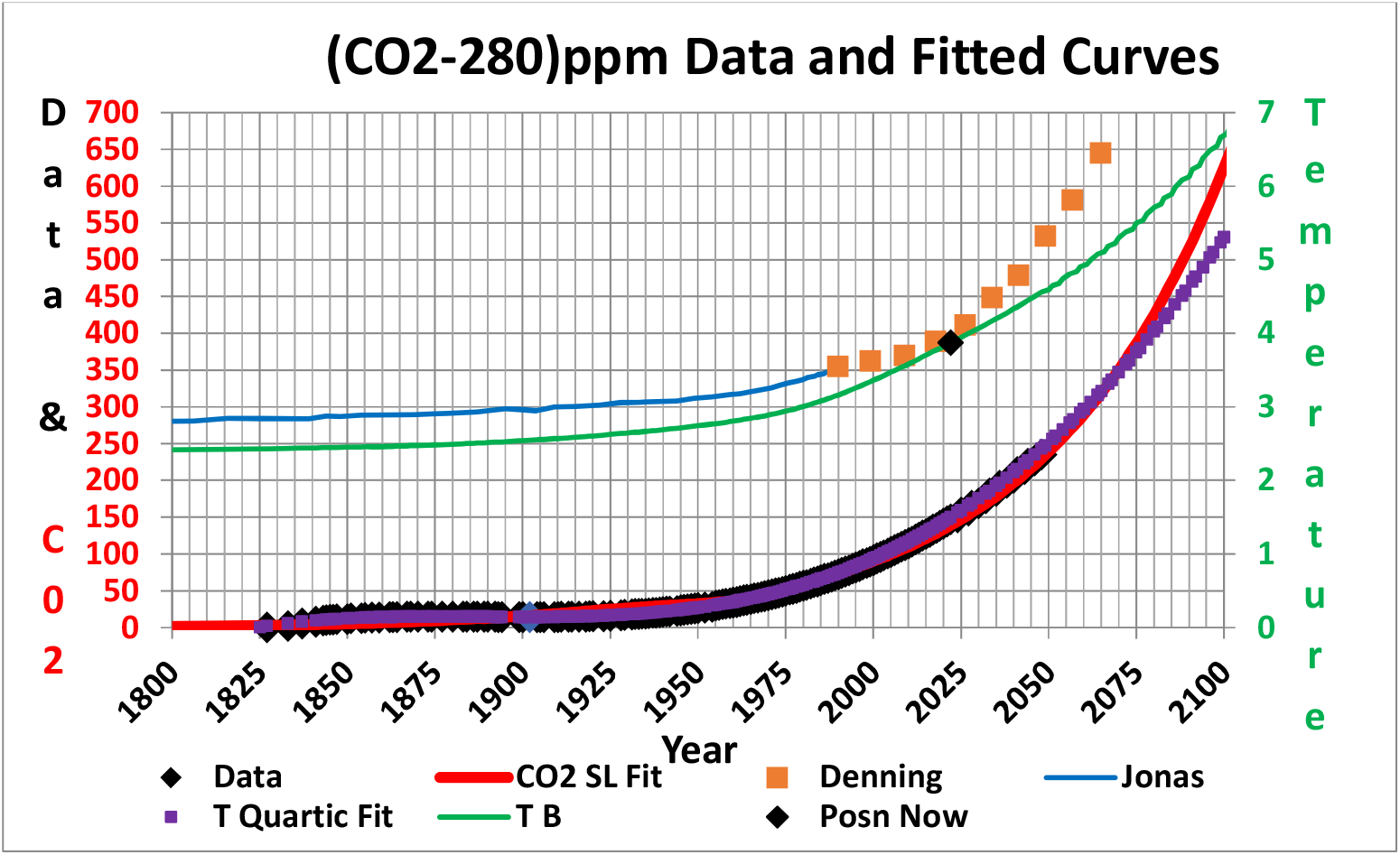
As in Figure 6(a). Also shown are the Jonas fit (the most optimistic and the Denning calculation. Also shown is the fit to the CO2 using the Quartic terms reflecting the Jonas calculation described also in Subsection 4.3.

## 5 ERROR ESTIMATION

### 5.1 Bootstrap Analysis

We use bootstrap analysis to calculate confidence levels of the fitted quantities. The method is well-known (see for example, Cheng 2017; Dye et al. 2020, Cheng et al. 2020), so full details are not repeated here.

In summary, to estimate confidence levels for *D*(*t*), we generate a parametric bootstrap sample *E*(*t*_*i*_) *i=* 1,2,…,*n*, where

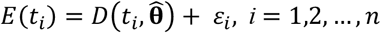

and 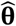is the ML estimate of **θ**, and

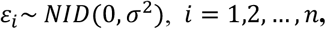

is a random sample of mutually independent distributed, normal pseudorandom variables with variance *σ*^2^. This is carried out *B* times so that we have

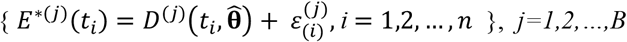

with the asterisk denoting that the observation is bootstrapped.

We can now estimate the parameters from each of the samples giving the bootstrap parameter estimates and bootstraps functions:

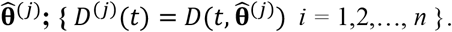

From these a confidence interval can be obtained for each parameter using the ranked values of that parameter.

A similar method can be carried out for *D*(*t*). Note however that this only gives the confidence interval level at each *t*_*i*_ separately. the confidence level is reduced if several different *t*_*i*_ are considered simultaneously. A more complicated method is needed using maximized likelihood regions. See Cheng (2017) for details. We could have used the simpler method in this paper, but made a modification to model the dependence of an observation on previous observations. Specifically the observations assumed to be a first order autoregressive process : Thus we set

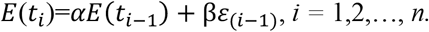

The standard deviation of an observation is therefore not *σ*, but is *s*, which is approximately

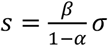

Thus values of *α* = 0.75, *β* = 0.25, would make *s* = *σ*.

In the experiments reported here we actually varied the value of *β* slightly to remove possible bias in the estimate of *σ*. This is not of great practical consequence as our results are only for discussion and are not used in practice. In any case our interest is centred on the SL parameters, and a well-known property is that the estimate of *σ* is asymptotically independent of the other parameters.

### 5.1 Covid 19 Bootstrap Results

Figure 7 illustrates the results of a representative set of three bootstrap scatterplots of pairs of parameters, obtained from *B* = 50 bootstraps. These indicate the typical scatter.

**Figure 7.**
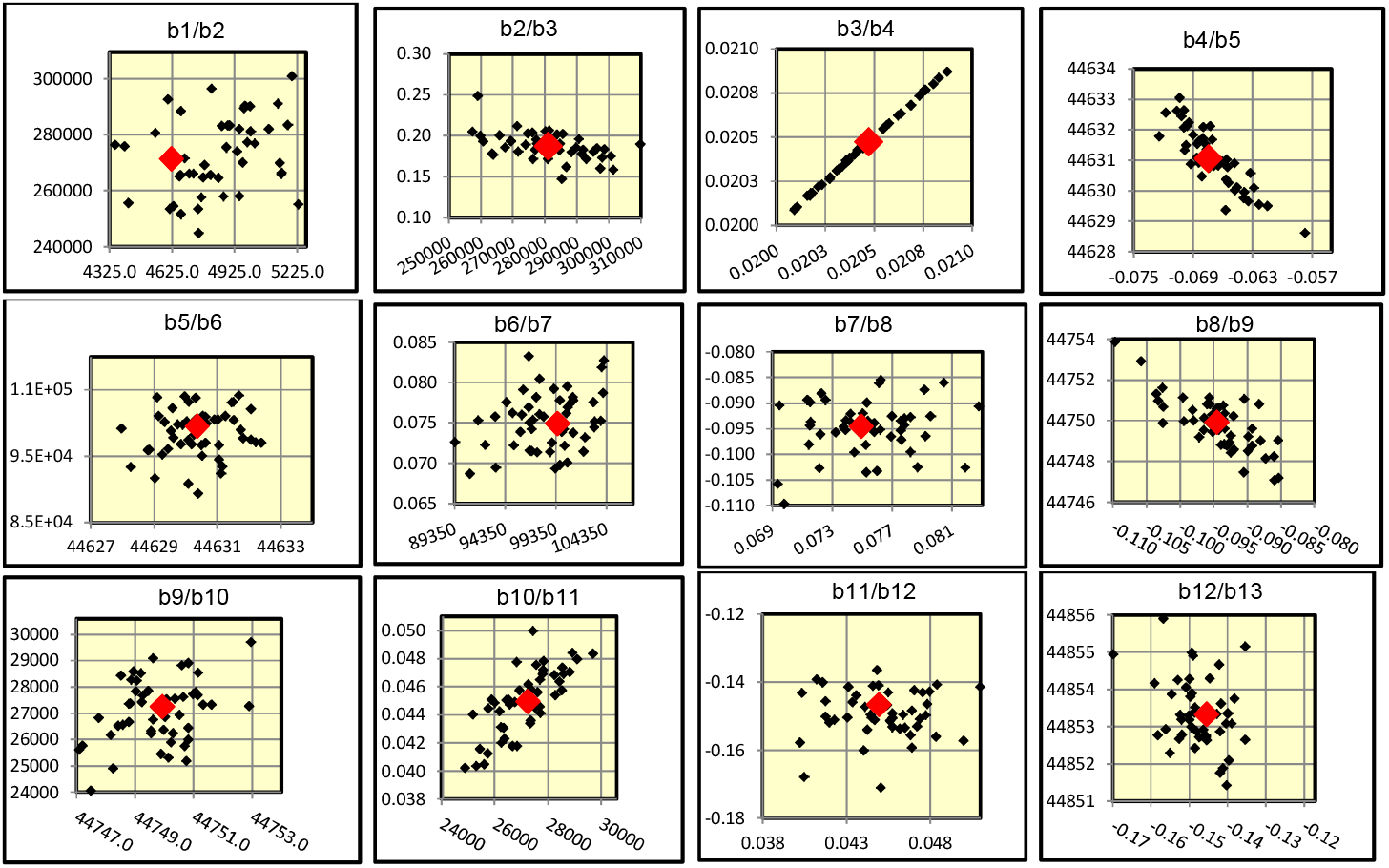
Some Bootstrap Scatterplots

Figure 8 gives shows the resulting *D*(*t*) plots of the MLE and the Upper and Lower CI curves with confidence level 90%.

**Figure 8.**
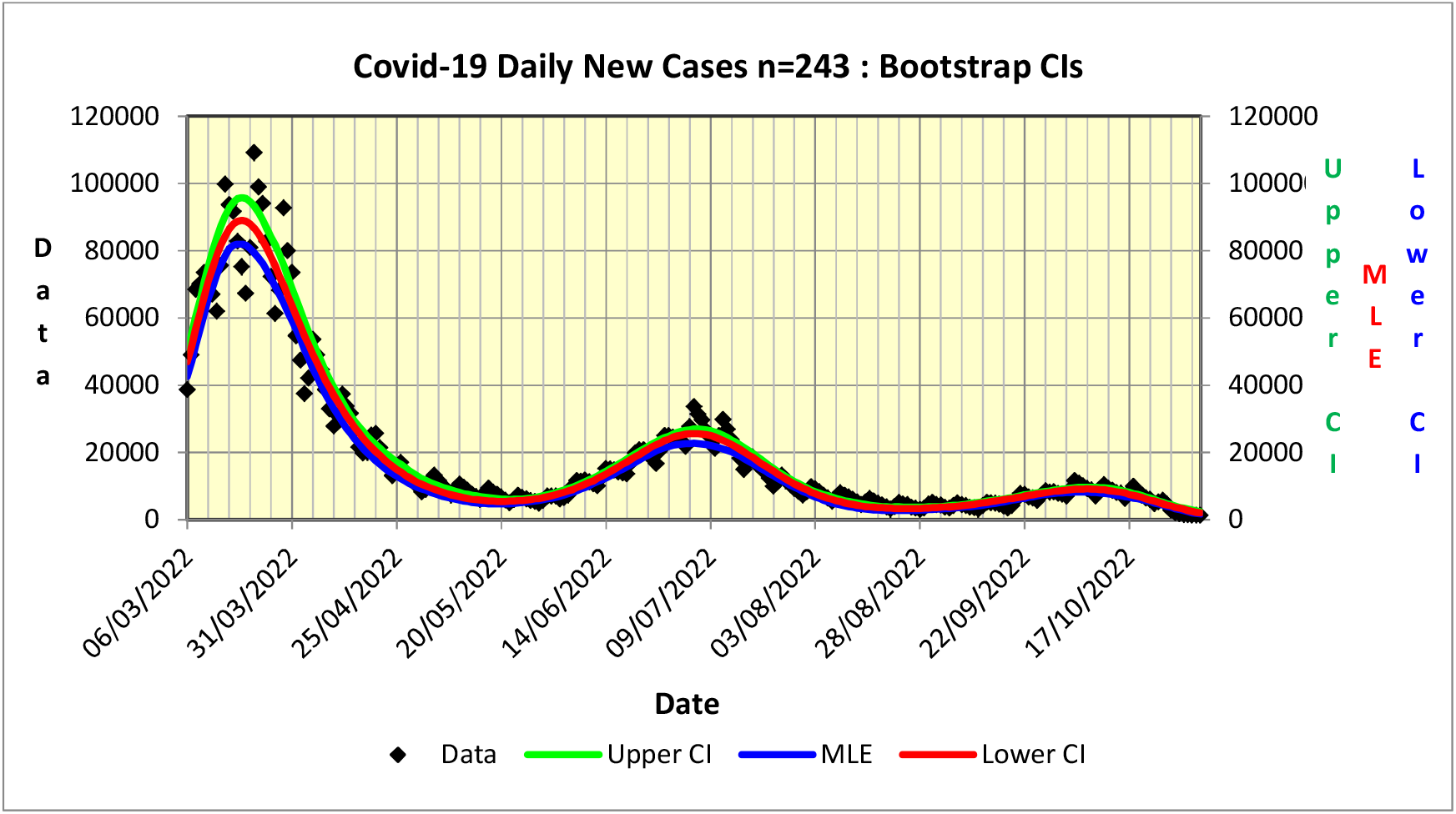
The N=3 Multi-wave fit to the COVID-19 sample

### 5.1 CO_2_ Bootstrap Results

Figure 9 illustrates the results of a representative set of eight bootstrap scatterplot pairs of parameters obtained from *B* = 50 bootstraps. These indicate the typical scatter.

**Figure 9.**
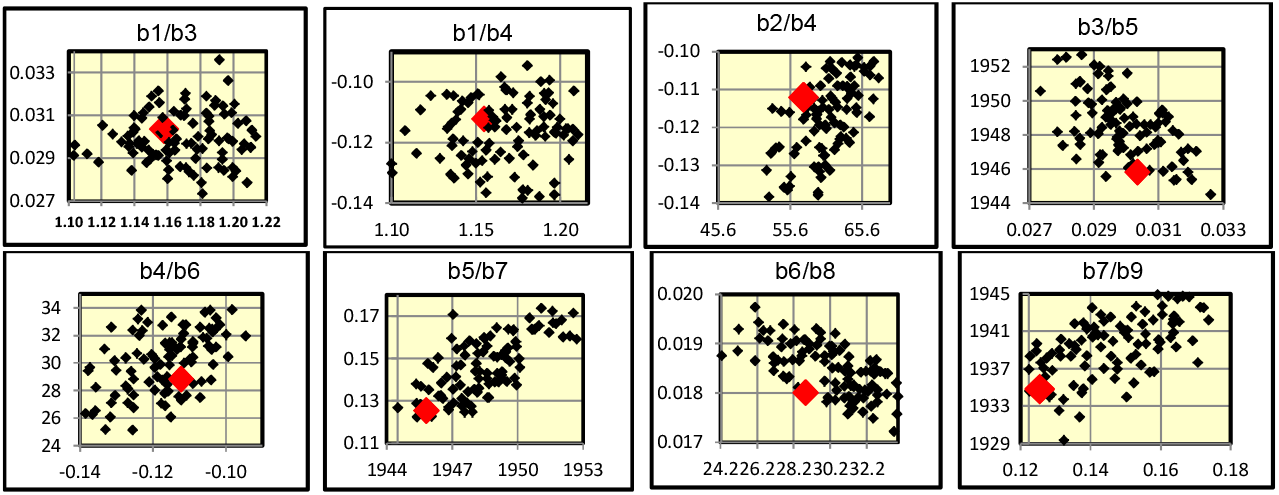
Some Scatterplots for CO_2_ Example

**Figure 10.**
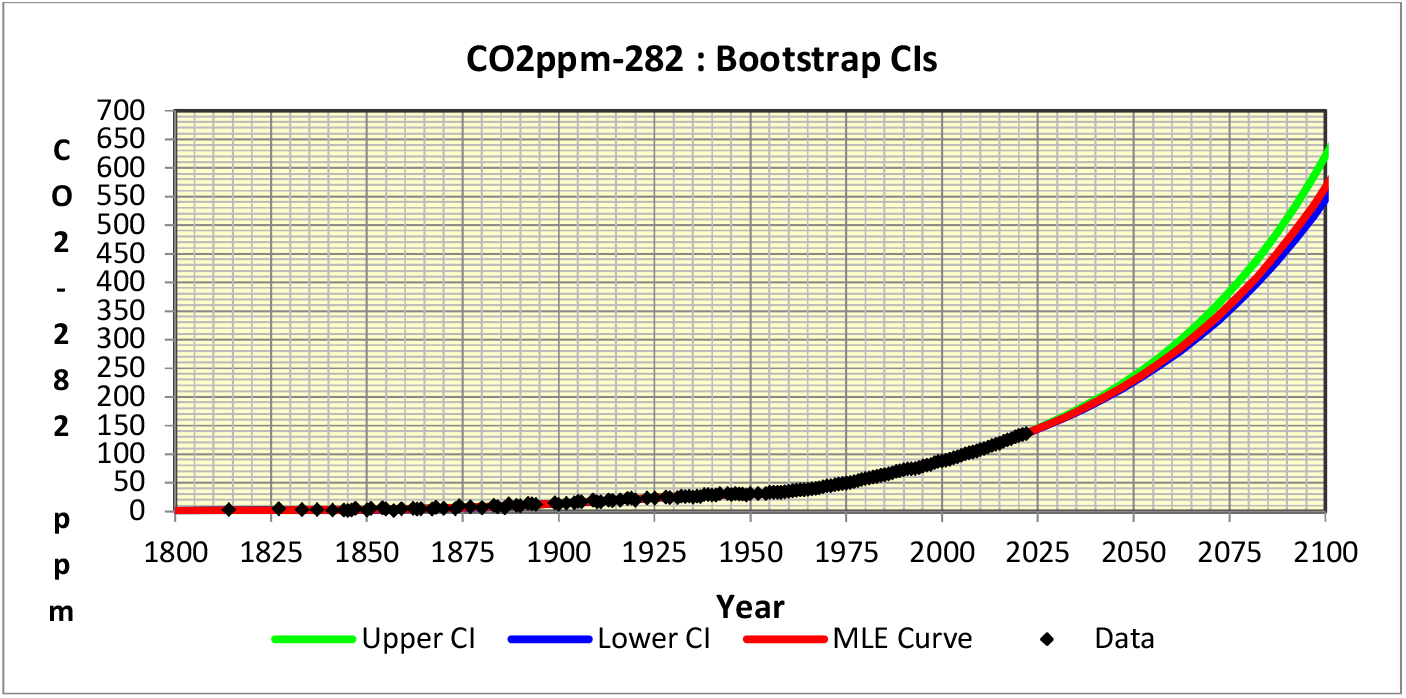
The N=2 Multi-wave fit to the CO_2_ sample

## 6 IMPLICATONS OF CO_2_ SCENARIO

To understand the impact that *Homo sapiens* has had on the earth’s climate it is necessary to deal with very large numbers. The weight of the earth is 5.97×10^24^ kg of which 9×10^19^ kg is carbon in the earth’s crust. The carbon in plants weighs 4.50×10^14^ kg very much less than in the earth’s crust. The carbon in 1 ppm CO_2_ in the atmosphere weighs 2.12×10^12^ kg. Before the industrial revolution, the concentration of CO_2_ in the atmosphere fluctuated, between about 190 ppm and 270 ppm for 800k years, with a period of about 100k years, while the rises tended to be faster than the declines. The weight of carbon in the atmosphere was therefore 4.87×10^14^ kg, about the same as the weight of carbon in plants, fluctuating from about 4.03×10^14^ to 5.72×10^14^ kg.

Since the industrial revolution the concentration of CO_2_ in the atmosphere is now about 415 ppm (Figure 6) as the result of burning fossil fuels, so that the weight of carbon has increased to 8.80×10^14^ kg or about twice weight of carbon in plants. The consequence of this is that the earth’s atmosphere has already experience an increase in temperature of about 2°C (Figure 6) with the prospect that, if the concentration of CO_2_ continues to increase, following the current trend, the temperature could increase by 6°C above preindustrial levels which would be catastrophic for our survival. We therefore need to sequester CO_2_ on a massive scale.

In order to return to pre-industrial levels of atmospheric carbon and therefore temperatures, we need to remove about 4×10^14^ kg of carbon from the atmosphere or 1.5×10^10^ kg every day from now until the year 2100. The present world population is 8 billion. This means, on an individual basis, we each have to ensure that 2kg of carbon is returned to Earth each day.

The biomass of rain forests is 500 metric tons per hectare or 50 kilograms/m^2^ so that one would have to expand the worlds rain forest, which currently occupies a 7 million square metres, by about 12k square meters every day until the end of the century. Efficient ways to sequester carbon from the atmosphere are desperately needed.

A recent discussion suggests that systematically scattering iron-rich dust onto target areas in oceans around the world could sequester perhaps 3×10^13^ kg carbon per year, or about 10^11^ kg per day, if the world’s deep oceans were to be treated annually. Using a new method of carbon capture it seems that one could remove carbon at a cost of US$0.5/kg of carbon which means that it would cost about 10^9^ US$ per day, every day from now until 2100 while the worlds GDP is 2.7×10^10^ per day. These studies suggest that it may be technically feasible to sequester carbon on the necessary scale but this would have to be done on an extraordinary scale and much more efficiently than is now possible.

The long term survival of our species depends on it.

## Data Availability

All data are available online.

https://doi.org/10.1098/rsos.201726

https://archive.ph/FC7zE

## AUTHOR BIOGRAPHIES

**RUSSELL CHENG** retired from the University of Southampton in 2007 where he had been Head of the Operational Research Group, having held previous positions at Cardiff University and the University of Kent at Canterbury. https://www.southampton.ac.uk/maths/about/staff/rchc.page

**BRIAN WILLIAMS** is Senior Research Fellow at the South African Centre for Epidemiological Modelling and Analysis (SACEMA) having held the position of Epidemiologist at the World Health Organization from which he retired in 2008.

